# Quality and safety for the public through domiciliary nursing in Italy: a multicentre cross-sectional descriptive observational study (The AIDOMUS-IT Protocol)

**DOI:** 10.1101/2022.12.06.22283098

**Authors:** Annamaria Bagnasco, Rosaria Alvaro, Loreto Lancia, Duilio Fiorenzo Manara, Laura Rasero, Gennaro Rocco, Zega Maurizio, Beatrice Mazzoleni, Loredana Sasso

**Affiliations:** Department of Health Sciences University of Genoa, Via A. Pastore, 1 16132 Genoa, Italy; Department of Biomedicine and Prevention, Faculty of Medicine, University of Rome Tor Vergata Via Montpellier, 1, 00133 Rome, Italy; Department of Clinical Medicine, Public Health, Life and Environmental Sciences, University of L’Aquila, Piazzale Salvatore Tommasi, 1, 67100 Coppito, L’Aquila, Italy; Vita-Salute San Raffaele University, Via Olgettina, 58, 20132 Milano; Department of Health Sciences, University of Florence, Florence, Italy; Centre of Excellence for Nursing Scholarship, c/o OPI Roma, Viale degli Ammiragli 67 sc. B, 00146 Rome, Italy; University Policlinic A. Gemelli IRCCS, Largo A. Gemelli, 8, 00168 Rome, Italy; Humanitas University, Via Rita Levi Montalcini 4, Pieve Emanuele, 20090, Milano, Italy; Department of Health Sciences, University of Genoa, Via A. Pastore, 1, 16132 Genoa, Italy

**Keywords:** family and community nurses, community care realities, complexity of care, multicentre observational study

## Abstract

**Introduction:** The development of this study protocol occurred in conjunction with the new Regulation defining models and standards for the development of community care in the National Health Service (Ministerial Decree n. 77-2022) with the redefinition of care in the community. Considered the increase in the elderly population, in the complexity of care and the workload for home care, it is important and urgent to describe the work environment and the Italian community care reality. The main objective of this study is to evaluate the characteristics of nursing care and the quality of home care in the community in Italy.

**Methods:** This is a cross-sectional descriptive observational study using the survey method. The study protocol uses quantitative data from three sources: 1) primary data on organizational characteristics, professional satisfaction, intention to leave home care services, and burnout; 2) secondary data on the experience of patients and their informal carers; 3) data on improper access to the emergency department, readmission to hospital, comorbidities, services offered, and user level of autonomy, and main and secondary diagnoses. Data will be analyzed using descriptive and inferential statistics.

**Conclusions:** The systematic analysis of the different national community care contexts will enable to increase the knowledge and awareness of the need for community health care. The availability of specific data will promote and support the role of the family and community nurse in district health care contexts, to increase the quality of care perceived by patients and their families.

## Introduction

In the literature, it has been shown that there is a clear association between the nurse-patient relationship and the patient mortality and/or worsening of health deterioration (Senek et al., 2020). The European study, RN4CAST, has shown that in hospitals with higher nurse staffing levels, there are better outcomes for both patients and nurses (Aiken et al., 2017). Although the main outcome measure was mortality rate, the RN4CAST study also enabled to investigate other indicators such as patient safety, adverse events and quality of care, nursing ratios and skill-mix (Senek et al., 2020). However, most of the studies concerning the nurse-patient ratios and the quality of care, have been conducted in hospital settings (Poghosyan et al., 2017; Poghosyan et al., 2019).

As the elderly population and chronic degenerative diseases increase, care and social complexity is greater, generating a growing demand for nursing care at home or “closer to home”, with particular attention to timely and appropriate discharge from hospital (Chapman et al., 2019; Poghosyan et al., 2017). This situation leads to a growing need for nursing care at home (Veenstra et al., 2020), with new and innovative health care and support approaches (Chapman et al., 2019). Moreover, with the reduction of hospital resources and the increase of hospital care costs, a greater percentage of health services are transferred from specialized hospital services to community care facilities (Spasova et al., 2018). This situation, in turn, entails increased responsibilities for home care nurses towards older people with increasingly complex care needs (Veenstra & Gautun, 2021) and nursing competencies to meet people’s needs.

Studies concerning the quality of nursing care in community settings mainly explore three areas of care: domiciliary care, nursing homes, and primary care or district nursing. Domiciliary care includes services provided in the patient’s home for the purpose of promoting, maintaining or restoring health or minimizing the effects of illness and disability (Möckli et al., 2021). Some studies, which identify common variables across multiple areas of care, tend to identify the community context with more than a single area (Coffey et al., 2019). There are many definitions for the quality of health and social services, and they frequently include concepts of effectiveness, timeliness, safety, equity, efficiency, and being patient centred. According to the analysis of Aase (2021), quality in community settings is understood by health professionals as pride and dignity in their professional practice. Quality is conceived as a continuing process that depends on having the “right competency” and ability to cooperate with other professional groups, placing the patient at the center of all the activities. The qualitative dimensions of the structural characteristics (i.e., system, documentation) and the “soft” dimensions (i.e., dignity, relationships) are important for the conceptualization of quality by health professionals and should be considered when conducting quality improvement activities and in daily practice. In community care settings, domiciliary care enables to take charge of patients and their care needs, allowing a better response in terms of outcomes (Möckli et al., 2021).

Approximately 14-25% of hospital readmissions and accesses to the Emergency Department (ED) of nursing home residents have been classified as potentially preventable (Unroe et al., 2018). In addition, readmissions and accesses to the ED are associated with increased healthcare costs, patient complications, healthcare-related infections, and other adverse consequences. Therefore, minimizing avoidable readmissions and accesses to ED is a top priority for the improvement of global health care (Yang et al., 2021). One of the causes of hospital readmission and inappropriate access to the ED are the interruptions in communication between home and local care staff, patients and families, and providers; flaws in healthcare processes; lack of resources including equipment, qualified staff, or on-site providers; and failure to identify home care goals (Unroe et al. 2018). Poor communication and organization between domiciliary and clinical nursing also undermines the quality of home care and leads to readmissions or improper accesses to the ED. Poor communication and organization also constitutes a major challenge for the quality of home care services, as it can lead to negative patient outcomes, poorer health conditions, and unnecessary or incorrect treatment resulting in waste of resources and increased healthcare costs (Möckli et al., 2021).

With greater workloads and increasing home care complexity, the need to provide appropriate staffing levels and skill-mix is a constant challenge (Chapman et al., 2019). Domiciliary care does not only involve patient health aspects related to post-hospitalization, but also other important aspects of care management, such as therapeutic education or patient follow-up (Poghosyan et al., 2017). Rationing care or not taking charge of patient care routinely, has potential safety implications for complex patients living in their homes, and increases the amount of time required to care for these patients (Sworn & Booth., 2020). In addition to the increasing complexity of care, domiciliary nurses often find it difficult to provide services due to a lack of resources to address the additional responsibility of having higher numbers of older patients with chronic illnesses discharged from hospitals often also inappropriately (Veenstra et al. 2020). At the clinical level, the phenomenon of missed care is often associated with problems linked to lack of time and staff, which forces nurses to ration care based on healthcare priorities (VanFossen et al., 2016).

In a study by Senek et al. (2020), conducted in the UK, it was shown that there was a relatively low prevalence of care left undone within the domiciliary/community nursing care setting. This could be due to the presence of appointment systems with clearly defined schedules compared to other domiciliary/community settings (Senek et al., 2020). In fact, home care is not continuous and includes daily or weekly visits, this situation often increases the workload of informal carers or family caregivers (Möckli et al., 2021). To meet the increasing complexity of home care, it is necessary to have sufficient staff to respond to care demands (Griffiths et al., 2016). The shortage of nursing staff in domiciliary/community health services is a recurring concern often reported in the literature (Veenstra et al., 2020). Low levels of nursing staff in domiciliary/community care settings are associated with negative home care experiences (Griffiths et al., 2016), increased hospital readmissions and less continuity of care (Veenstra & Gautun, 2021).

One of the problems of domiciliary/community nursing care is the absence of a single staff-patient relationship that can be applied to the full range of domiciliary/community care services to ensure safety and meet patients’ healthcare needs (Veenstra et al., 2020). In addition, little evidence is available regarding what constitutes an appropriate level of nurse staffing in various community settings, making it difficult to implement staffing requirements or legislation in the community setting (Senek et al., 2020). Baker et al., (2018) highlighted how the absence of a home nursing coordinator negatively affects the outcomes of domiciliary care, leading to increased hospital readmissions.

In addition to understaffing, another issue is the insufficiency of skill-mix required to provide high quality domiciliary/community care (Veenstra et al., 2020). Care at home is provided by various health professionals, including physicians and nurses, whose scopes of practice often overlap by working in teams that have a variable and less defined organizational structure than those in specialized outpatient and hospital settings (Poghosyan et al., 2017). Having the right number of properly trained, competent, and experienced nurses is important to achieve better patient care outcomes, reduced mortality rates and increased productivity (Sworn & Booth, 2020). Furthermore, domiciliary nurses not only interact with other health professionals of the team, but also with informal carers who take care of people at home or with other health professionals who directly or indirectly take part in the care provided (e.g., health care workers and general practitioners) (Möckli et al., 2021).

With the new “Regulation defining the models and standards for the development of community care within the National Health Service” (Ministerial Decree No. 77/2022) with the redefinition of community care and the inclusion of a greater number of family or community nurses on the Italian territory, it is timely and important to evaluate the characteristics of nursing community care in Italy.

## Aims

### Main objective

To evaluate the characteristics of nursing community care management and the quality of home care in Italy.

### Secondary objectives

- Describe the staffing levels in the field of domiciliary nursing.
- Describe the levels of work environment in the field of domiciliary nursing.
- Describe the levels of caseload in the field of domiciliary nursing.
- Describe the phenomenon of missed care in the field of domiciliary nursing.
- Identify the levels of skill mix for the quality of care in the field of domiciliary nursing.
- Describe the level of quality perceived by home care recipients and their caregivers.

## Methods

### Study design

This is a multicentre cross-sectional descriptive observational study that uses the survey method. The study uses quantitative data from three sources:

1. Collection of primary data through an online questionnaire sent to domiciliary nurses at a single point in time. This source will enable to collect data on organizational characteristics (e.g., nursing work environment). In addition, also data regarding nurses’ job satisfaction, intention to leave home care, and burnout will be collected.
2. Secondary data on the experience of patients and their informal carers. This source will enable to collect data relating to the way their care is managed, their communication with health professionals, and between health professionals, information received while home care is being provided, information received on how to manage their care at home after the intervention at home.
3. detailed data from home care clinical-healthcare databases with a special focus on improper accesses to the emergency department, readmissions to hospital, comorbidities, services offered, and care recipients’ level of autonomy, and main and secondary diagnoses (ministerial indicators of the information system for monitoring domiciliary care [SIAD]; indicators of the New Guarantee System for monitoring health care – inter-ministerial decree of the 12^th^ of March 2019 art.3, paragraph 1).

### Study context

The study will be conducted on home care services provided in community districts at a national level. Home care is a service provided in the district aimed at providing home-based interventions characterized by different levels of intensity and complexity including specific care pathways and personalized care plans.

### Study population

The target population for this study are home care nurses, home care recipients and their informal carers (for the purposes of this study, the informal carer is the contact person identified by the healthcare professionals of the districts). Participation implies acceptance of the letter of presentation of the study by the General Director of the community healthcare centre.

### Eligibility criteria

Inclusion criteria for the nurses:

- Being a registered nurse
- Provide domiciliary care in a district of the Italian territory
- Provide informed consent

Inclusion criteria for care recipients / informal carers:

- Care recipients and their informal carers taken charge by the Community services participating in this study.
- Informed consent for care recipients and their informal carers

Exclusion criteria for the nurses:

- There are no exclusion criteria regarding age, sex, ethnic group or socioeconomic status. There are no enrolment restrictions in relation to fertile status, pregnancy and/or other nurses’ characteristics.

Exclusion criteria for care recipients/informal carers:

- Care recipients and informal carers who access community services, but also use other services that do not involve domiciliary care (e.g., outpatients’ services).

### Study variables

- Staffing and skill mix of the nursing team in community care, defined in terms of hours of care provided to each patient, nurse-patient ratios, or an equivalent measure (e.g., nurses per district population - NHS, 2022)
- Work environment, defined as the organizational characteristics of a work environment that facilitate or limit professional nursing practice (Lake et al., 2002)
- Caseload, defined as the set of activities provided to people who require care from the local nursing service in a specific period and in a specified location (NHS, 2022)
- Missed Care, defined as any aspect of the care needed by a person that is omitted (in part or totally) or delayed (Kalisch et al., 2009)
- Skill Mix, defined as the proportion of nurses compared to total care staff, including nurses, general nurses, and health care workers in each unit (Kalisch & Lee, 2012)
- Customer satisfaction, defined as the level of the satisfaction of care recipients and their informal carers with the care provided by health professionals (Manzoor et al., 2019)

### Procedures

The Nursing Managers / Contact persons will be invited to participate in organizational meetings with the Scientific Director and/or the members of the Scientific Committee. In addition, they will be asked to provide data on the population demographics and characteristics, the characteristics of the territory (i.e., size, services offered), staffing, and the types of services provided.

All the nurses of the participating centres will be invited to participate via e-mail, which will contain a link enabling a reserved access to the information material and the study questionnaire. By responding to the completed questionnaire, the participants confirm their consent to participate in the study.

For the respondents (i.e., nurses, care recipients, and informal carers), the duration of their participation in the study will be short and the questionnaire will be administered only once. No other study procedures shall require a follow-up with the participants.

### Instruments

#### Sample size

The minimum sample size required for each university is 460 nurses, based on a 5% margin of error, a 20% dropout rate, and a 95% level of confidence.

Considering the total Italian nursing population of approximately 450,000 on the register of the Orders of the Nursing Professions, a sample size of approximately 1% of the population or 4,600 nurses was therefore hypothesized.

### The Nurse Survey

The first page of the questionnaire (Appendix 3) provides information about informed consent, including the purpose of the study, assuring participants that their responses will be kept confidential and not disclosed, especially to their employer, the voluntary nature of the study, a webpage with frequently asked questions (Appendix 4), and the contact information of the PI for Italy and the PI of the participating center. By completing and sending the questionnaire participants express their consent to participate in the research.

The data collected through the questionnaire be entered into a secure electronic platform kept by CERSI-FNOPI, independent from the participating hospitals and approved for research purposes of this kind:

a. The coordinators of the participating community districts will send a weekly reminder via e-mail or WhatsApp to solicit participation in the study.
b. No identifying information will be required from interviewees as part of the questionnaire ensuring the confidentiality of the answers and will be unknown even by the research group.
c. In case nurses prefer to answer questions outside their work environment to protect the confidentiality of their answers, the questionnaire can be completed on any device: personal smartphones, tablets, or PCs at home. The questionnaires are compatible with all web browsers.
d. Partly completed questionnaires that have been suspended can be resumed where the respondent left off without having to start all over again.
e. User-friendly dashboards will enable to provide real-time response rates to the research team, which will be communicated to the PI of the participating center.

The survey data will upload directly onto the CERSI.FNOPI.IT server. The community healthcare centres will receive their data in the form of a report on the results of their center. The information will be aggregated to protect respondents’ confidentiality.

An e-mail address and telephone number will be provided on each communication sheet and page of the electronic survey, which respondents may use to solve technical problems or ask further information about the questions.

After finishing the survey, the anonymous data of the survey responses will be exported and saved on password-protected servers hosted by a certified service provider. The original anonymous raw data will be kept by CERSI-FNOPI for ten years.

### The Patient Survey

Information on the patients’ outcomes will be drawn from the data in the medical and nursing records regarding re-hospitalizations, comorbidities, services offered, autonomy level of the care recipient and principal and secondary diagnoses that will be collected by the PI of the participating center.

Patient satisfaction data will be derived from the patient survey. Patients will be recruited by home care nurses based on the inclusion criteria. The first page of the survey (Appendix 5) provides information on informed consent, including the purpose of the study, assuring participants the confidentiality of their responses, the voluntary nature of the study and the contact information of the PI for Italy and the PI of the participating center. The informed consent form must be signed by the patient before participating in the study.

Information on patient outcomes derived from data in hospital databases regarding improper accesses to the Emergency Department and re-hospitalizations will be collected by the PIs of the participating hospitals or their delegates.

The selection procedure will be evaluated based on available data regarding the organization of services, obtained from the exploratory survey described in phase two of the study.

### The Informal Carer Survey

Informal carer satisfaction data will be collected through the Informal Carer Survey. The first page of the survey (Appendix 5) provides information on informed consent, including the purpose of the study, assuring participants the confidentiality of their responses, the voluntary nature of the study and the contact information of the PI for Italy and the PI of the participating center. The informed consent form must be signed by the informal carer before participating in the study.

### Benchmarking procedures

The community healthcare centres can choose to receive a final benchmarking report that compares each centre with the anonymised data of the other centres. To protect the confidentiality of participating nurses, community healthcare centres will receive a report on their centre’s results only in an aggregate form.

The report will be shared with the PI of the participating center.

### Data analyses

The preliminary analysis of specific raw data will be conducted to detect any inconsistent and missing data. The clean data will be divided into datasets containing information at various levels: district/zone/community, at home, patients/informal carers, and nurses.

Two main analyses will be conducted: the first will be a descriptive and comparative analysis of the variables that summarize similarities and differences, and strengths and weaknesses of the nursing workforce. In these analyses, data at the nurse level will be used in an aggregate form.

The second will use models for the detailed analysis of the associations between independent and dependent variables. The analysed independent variables will concern staffing and variables regarding the work environment. The dependent variables will include both the indicators of nurses’ perception of work and patient outcome variables.

Analyses will be conducted using regression models to estimate the mean differences for the continuous outcome variables or differences in the odds ratios of various negative events for nurses and patients.

Multivariate analyses will be conducted for all nurses and patients in the individual districts/zones/communities. Three analyses will be conducted, the first will explore factors related to nurses’ work environment with procedures for risk adjustment. Logistic regression models will be used with data on patient caseload (demographic characteristics, comorbidities, diagnostic categories, level of autonomy) to identify the factors that most influence the quality of home care.

The second analysis will evaluate the association between the characteristics of the district/zone/community (workloads, and work environment) and nurse outcome measures.

The third analysis will evaluate the association between the characteristics of the district/zone/community (workloads, and work environment) and the patient outcomes (data collected from the clinical records of the home care services in the community).

The fourth analysis will evaluate the economic impact in terms of costs and outcomes, by analysing the direct and indirect costs, and other outcomes of concern as envisioned by the National Health Service. An impact analysis on the five-year budget will be prepared for the proposal of alternative strategies.

### Ethical considerations and permissions

This study protocol was developed in full respect of the rights of the participants by abiding to the ethics and conduct required by Good Clinical and Research Practice (Ministerial Decree 14/7/97), although no medications will be administered and according to the General Data Protection Regulation (GDPR) EU Regulation 2016/679.

The study will be conducted in agreement with the Declaration of Helsinki (Fortaleza 2013 version) and current rules on clinical trials and good clinical practice.

This study requires that all participants provide in informed consent in each phase of the study by reading the comprehensive “Information Sheet” online with the necessary information that participants must be aware of.

The information sheet can be read when opening the link to complete the online questionnaire. After reading it, participants may give their consent or not to participate, and the section with the questions can be accessed only after confirming their consent.

### Approval of the protocol

Since personal questions and socio-personal information will be collected, before starting the study, the protocol will be submitted to the Liguria Regional Ethics Committee. If the other participating centres require the approval of their own local ethics committees, all the necessary documentation will be sent.

### Ethical aspects and confidentiality

The promoter of this study shall protect sensitive personal data, both clinical and non-clinical, of the subjects involved in the study in agreement with European legislation (EU Regulation GDPR 2016/679). Absolute voluntary participation in the study and the decision to refuse consent or withdraw from the study at any time during its execution will be guaranteed.

### Confidentiality and warranty rules

All information collected for this study will be anonymous and will be entered onto a database and processed according to the present protocol and in compliance with the current privacy regulations. All collected data will only be made available to the research team. Any breach of data security will be reported to the relevant authorities within 48 hours. To protect privacy, the data of individuals will be entered through an online system with reserved and controlled access, using a numerical identification code standing for the Research Center and an individual progressive number standing for each individual. The participants’ initials, date of birth, or other personal information which may enable to track their identity will not be recorded.

This study involves the use of an online data collection form, prepared by the Promoters and the Scientific Committee of the study. The form was designed to facilitate its compilation and to make data immediately available anonymously to the Study Promoter, to allow both a priori quality control of the data entry (i.e., through appropriate filters that block the insertion of inconsistent information) and a remote verification of the data entered. Access to the data collection form is password protected. The data of the subjects will be saved in the database anonymously and only the investigators shall be able to track the identity of the individual.

The researchers involved in this study undertake to respect the confidentiality of specific data collected and analyzed for each individual institution (including a commitment not to disclose or publish specific data of any entity in any identifiable way). The anonymized results will be read and commented only by the members of the research team. The data, made totally anonymous, will be kept for at least ten years by CERSI-FNOPI. In addition, each participant will be given the opportunity to express doubts and any requests for clarification regarding the study in progress, by providing the contact information of the research team. It is expected that there may be benefits for those taking part in the research, in terms of gaining a deeper understanding of this emerging phenomenon in the international scientific literature. Participants shall not receive any fees or reimbursements, at any level.

## Discussion and conclusions

In the literature, the importance of nursing care in community settings is widely discussed and how the family and community nurses can contribute to reduce public health expenditure and re-hospitalizations (Spasova et al., 2018). Community nursing is widely studied in other countries, and its importance has been highlighted also during the COVID-19 pandemic (Goh et al., 2021; Shapiro et al., 2021). Community nursing care enables to take charge of patients and support their families or informal carers by implementing innovative healthcare approaches (Chapman et al., 2019).

Due to the increasing older population, workload of domiciliary community care and the increase in care complexity (Veenstra & Gautun, 2021), it is important to have the capacity to describe the work environment and the community care reality in Italy. It is necessary to systematically analyse the various national community care contexts, as proposed in this study protocol, to increase the knowledge and awareness of the community healthcare needs.

The availability of specific data will promote and support the role of the family and community nurse in territorial health care contexts, to increase the quality of care perceived by patients and their families. The study funded by CERSI-FNOPI is the first national project of the Center of Excellence for Nursing Research and Development (CERSI) in collaboration with nursing researchers and full professors of nursing of the Med/45 Sector - Nursing Sciences - for a research project led by nurses for nurses.

The development of this study occurred in conjunction with the new “Regulation defining models and standards for the development of community care in the National Health Service” (Ministerial Decree no. 77/2022) with the redefinition of care in the community. The CERSI scientific committee decided that this is the right time to collect descriptive data starting from the opinions of domiciliary care nurses and by asking Italian nurses to dedicate part of their time to participate in the study and give a voice to nurses through FNOPI.

For these reasons it was deemed necessary to launch the study on home nursing care in Italy and to highlight the current characteristics of home care nursing and the quality of care perceived by patients and their families.

The limitations of the study may be related to the number of missing answers, self-selection bias, and the organizational structure of home care in Italy, which differs across regions.

## Data Availability

All data produced in the present work are contained in the manuscript

